# Protocol for a randomized pilot trial of COMPASS, an open-source, culturally adapted cognitive behavioral therapy program for forcibly displaced Venezuelan adults in Peru

**DOI:** 10.64898/2026.03.19.26348265

**Authors:** Haley A. Carroll, Talia Guevara, Paula Gamarra, Chombalelo Mukunta, Shannon Dorsey, Bizu Gelaye, David Henderson, Matthew D. Bird, L. Feline Frier

## Abstract

Task-sharing approaches have shown promise in low-resource settings, yet few culturally adapted interventions have been systematically evaluated for forcibly displaced populations. Since 2016, over 1.7 million Venezuelans have migrated to Peru, facing significant barriers to healthcare and elevated risks of anxiety, depression, and post-traumatic stress disorder (PTSD). This protocol describes COMPASS (Cognitive-behavioral Open-source Mental-health Program Adapted for migrants, Sustainably delivered by lay providers and Supported by evidence). COMPASS is a transdiagnostic, open-source cognitive behavioral therapy program co-designed with forcibly displaced populations. This protocol describes the procedures for an ongoing randomized pilot trial with n = 90 forcibly displaced Venezuelan people (Clinicaltrials.gov: NCT06635486). COMPASS guides, or lay providers, trained through an intensive apprenticeship model, will deliver 6–12 weekly remote sessions. Primary outcomes include changes in anxiety, depression, and PTSD symptoms, assessed with validated Spanish-language measures. Secondary outcomes include feasibility (recruitment, retention, fidelity) and acceptability (therapist and participant ratings). Exploratory outcomes will examine integration, migration experiences, and demographic moderators of intervention effectiveness. Analyses will follow the intention-to-treat principle, using descriptive statistics and regression models to evaluate symptom trajectories across baseline, post-intervention, and 3- and 6-month follow-ups. This study represents the first effectiveness evaluation of an open-source, lay-delivered CBT program tailored for forcibly displaced people in Peru. Findings will inform feasibility, acceptability, and preliminary effectiveness of COMPASS, with potential to expand scalable, culturally relevant mental health services for forcibly displaced populations in resource-constrained settings worldwide.

## 1. Introduction

Over the past ten years, the global population of forcibly displaced people has nearly doubled, reaching 123 million^1^. While relationship between migration and mental health is a complex one, dependent on many factors such as events experienced prior to leaving, context of reception, or phase of migration, in metanalalytic research suggests that forced displacement, as opposed to voluntary migration, is a risk factor for mental health disorders like depression, anxiety, and post-traumatic stress disorders^2^. Additionally, approximately three-quarters of displaced individuals relocate to low- and middle-income countries (LMICs)^3^, where health systems are typically overburdened and barriers to care are substantial^4^. This highlights the need for scalable early interventions that can support the mental health of forcibly displaced people in LMIC settings.

Task sharing has emerged as a credible and pragmatic strategy for scaling mental health care in low-resource settings^5^, such as LMIC^6^. Evidence from brief, lay-delivered psychological interventions^7,8^ demonstrates clinically meaningful effects and feasibility in adversity-exposed populations. These approaches have shown promise even in highly vulnerable contexts, including among forcibly displaced people^8^, underscoring the potential of non-specialist delivery models to expand access to care where professional resources are scarce.

As one example of forcible displacement, since 2016, over 1.7 million Venezuelans have been forcibly displaced to Peru due to Venezuela’s humanitarian crisis, marked by extreme inflation, shortages of essential goods, and political violence^9^. Studies reveal significant mental health needs among these forcibly displaced people, where in transit 19% of forcibly displaced Venezuelans met criteria for depression and 23% for anxiety^10^. In Peru, forcibly displaced people face many legal, economic, and social barriers that hinder help-seeking, where they rely on shelters or community leaders for psychosocial support due to limited access to Peru’s formal healthcare system^4^. Although Peruvian law guarantees healthcare access to all migrants, including forcibly displaced Venezuelans, bureaucratic barriers such as the requirement for official documents limit access, exacerbated by deficiencies in Peru’s mental health infrastructure.

Against this backdrop, we developed COMPASS (Cognitive-behavioral Open-source Mental-health Program Adapted for migrants, Sustainably delivered by lay providers and Supported by evidence)^11^ guided by the Hueristic Framework for Cultural Adaptation^12^, the Ecological Validity Model^13^, the Taxonomy of Treatment Components^14^, and grounded in lived-experience. COMPASS was co-designed in partnership with forcibly displaced Venezuelans to directly address the acute shortage of mental health services in Peru. The program is open-source and delivered by trained lay providers, who are called guides, ensuring accessibility and scalability while maintaining fidelity to evidence-based practices. Its therapeutic content draws on a robust evidence base and incorporates core cognitive-behavioral components: behavioral activation^15^, cognitive restructuring^16^, problem-solving^17^, in-vivo^18^ and imaginal^19^ exposure, and emotion regulation^20^. COMPASS is not tailored to a specific mental health diagnosis, but is rather a transdiagnostic treatment protocol^21^, which targets the underlying and common features across mental health disorders with the primary targets being features of anxiety, depression, and post-traumatic stress disorder (PTSD).

Importantly, these skills are contextualized within migration-specific adaptation strategies, integrating culturally salient coping mechanisms co-developed with forcibly displaced communities (e.g., *pa’lante* and perseverance). In qualitative research we conducted with forcibly displaced people and experts^22^, we identified several distinct challenges faced by this group in their integration journey. These challenges included experiences of migration grief stemming from loss, discrimination, and xenophobia, as well as difficulties in reconciling their former “Venezuelan” identity with a new “Peruvian” identity. Ultimately, the interviews highlighted an urgent need for culturally sensitive, open-source CBT interventions tailored for forcibly displaced Venezuelan people in Peru, emphasizing essential themes such as cultural relevance, language access, and community involvement, while also addressing barriers related to integration and acculturative stress to improve mental health support for this community. Thus, a novel migration adaptation module was developed in collaboration with forcibly displaced people and experts. This module is optional, administered at the end of the program, and allows participants to apply skills they have learned throughout the intervention thus far.

Transdiagnostic and multi-problem treatment protocols are well established as effective, flexible, and sustainable options for mental health treatment in diverse populations. For instance, the Common Elements Treatment Approach^7^ utilizes common elements to target depression, anxiety, PTSD, substance use and other problems via lay providers^7^. The Common Elements Treatment Approach has been used with promising results in various global settings including with Bhurmese migrants in Thailand^23^ and in Somalian refugee camps^24^. The Unified Protocol is another transdiagnostic treatment option which utilize elements of altering thoughts, preventing avoidance of emotions, and helping take actions that are helpful to target the overlapping underlying origin of major emotional disorders like depression, anxiety and other related disorders^25^. The Unified Protocol has been modified for internally displaced people in Colombia to target depression, anxietym and PTSD as a result of armed conflict^26^. Additionally, culturally adapted CBT^27^ and culturally adapted CBT+ Problem management^28^ culturally adapts CBT to the population and uses emotional exposure and emotion regulation skills found in stretching, psychoeducation, meditation in various populations, including Farci speaking refugees^28^. However, COMPASS was designed over alternative existing treatment options because it uniquely combines open-source accessibility (i.e., eliminating licensing and training bottlenecks), migration-specific adaptation, utility with common mental health disorders (i.e., more skills and higher acuity of care than prevention or problem-solving options) and lay delivery. This design preserves the active therapeutic ingredients while enabling scale in resource-constrained environments.

A critical dimension of COMPASS is its emphasis on lived-experience involvement. Traditional implementation efforts often engage affected populations superficially^29^, without genuine shared decision-making, which limits effectiveness and sustainability. In contrast, COMPASS is grounded in the principle of “nothing about us, without us^30^,” ensuring that displaced and vulnerable populations are central to design and delivery^31^. This participatory approach enhances contextual relevance, fosters community ownership, and strengthens the likelihood of meaningful impact.

Thus, this study tests the preliminary effectiveness, and initial indicators of feasibility and acceptability of COMPASS via a pilot randomized control trial. This intervention addresses the mental health challenges faced by forcibly displaced Venezuelans living in Lima, Peru, focusing on common mental health disorders (CMDs) such as anxiety, depression, and PTSD. This study employs a waitlist control design to measure intervention effectiveness, hypothesizing reduced depression, anxiety, and PTSD symptoms in the intervention group over time. This study also seeks to expand knowledge on lay-provided mental health interventions for displaced populations in resource-scarce settings. If proven effective, this intervention could serve as a critical resource for community leaders, NGOs, and primary healthcare providers, addressing a pressing mental health gap.

## 2. Materials and Methods

### 2.1 Aim, Design and Setting of the Study

#### 2.1a Aim and Design of the Study

This is a randomized pilot study (Version 1.0, March 2026) with a waitlist control (WLC) which aims to compare trends in outcomes between the intervention (n = 60) and control group (n = 30). Participants are forcibly displaced Venezuelan people who have arrived and settled in Lima, Peru since 2014. Participants will be randomized via RedCap via allocated randomization set to assign 60 participants to the intervention group, and 30 participants to the control group. The allocation will be set by a member of the research team, and researchers will be blind to the sequence. After randomization, it will not be possible to keep the participant condition blind to researchers and participants, but lay person providers will not be informed to participant condition as intervention or WLC.

#### 2.1b Setting of the Study

This study takes place in Peru, with forcibly displaced Venezuelan people. According to Peruvian Migration Law, migrants have the right to access health services regardless of their status, a right that should be upheld by the Ministry of Health^32^. However, practical barriers exist, as many forcibly displaced Venezuelan people lack the foreigner or immigration card required to enroll in health insurance and access services. Additionally, the Peruvian mental health system faces significant deficiencies and inequalities, even for citizens^4^. Since 2007, a mental health reform has aimed to strengthen primary and secondary care, focusing on capacity building for the detection and initial treatment of mental health disorders in health centers and establishing community mental health centers for outpatient services^4^. Initially, Venezuelans arriving in the city were met with empathy. However, as the influx of migrants continued, perceptions shifted, and levels of xenophobia and discrimination increased related to migration status, socio-economic status, race, and gender^33^.

### 2.2 Sample size and Recruitment

Following the intent-to-treat principle, all randomized subjects will be included in the analyses. The sample size for pilot participants is based on a simple method outlined by Viechtbauer^22^, which suggests a sample size of n = 60 will be adequate for problem detection. As no standard treatment exists for forcibly displaced populations living in Peru, we will randomize n = 30 to waitlist control and treat all participants.

Participants will be recruited via engagement with NGOs active in migration, health centers and community mental health centers from different areas of the city, shelters that receive migrants, and schools. With permission we will distribute posters and fliers. Additionally, we will ask organizations to directly reach out to individuals they believe might be eligible to provide information about the study and give them the opportunity to learn more by a follow-up call by the research team. After completing the study, we will ask participants to notify their acquaintances who might be interested in the study and let them know that they can contact the research team directly. We will titrate recruitment based on the available number of open slots based on the number of lay person providers, the length of the intervention, and so forth. We will follow these recruitment procedures until our target recruitment is reached.

### 2.3 Inclusion and Exclusion Criteria

Inclusion criteria include qualifying for all of the following: (a) being an adult (>= 18 years old), (b) having Venezuelan nationality and arriving in Lima no earlier than 2014, and (c) symptoms of common mental health disorders including anxiety, depression and/or PTSD. Participants are considered to have a provisional diagnosis of: 1) anxiety if they score 10 or higher on the 7-item validated Spanish language Generalized Anxiety Disorder (GAD7) measure^34,35^, 2) depression if they score 10 or higher on the 9-item validated Spanish language Patient Health Questionnaire (PHQ-9) measure^36–38^, or 3) PTSD if they score 31 or higher on the validated Spanish language PTSD Civilian Checklist (PCL-5) measure^39^ with a criterion A event listed from the Lifetime Events Checklist (LEC5) measure^40^. Participants will not be restricted from pursuing other necessary care or services as recommended by other providers, NGOs, or other entities. Exclusion criteria include any of the following: (a) endorsement of suicidal ideation during screening, (b) endorsement of homicidal ideation during screening, (c) lifetime psychosis diagnosis, or (d) endorsement of problematic consumption of alcohol and/or other substances.

### 2.4 Description of the Intervention

#### 2.4a COMPASS Intervention

All participants will receive the COMPASS intervention, delivered by lay person COMPASS guides. COMPASS offers a flexible, transdiagnostic framework that incorporates evidence-based strategies which have demonstrated efficacy across various populations, conditions, formats, and contexts^41^ to tackle a variety of mental health challenges, including depression, anxiety, and PTSD. This structure enables the use of behavioral activation, cognitive restructuring, problem-solving, in-vivo and imaginal exposure, and emotion regulation treatment components to align with the individual needs of clients, making it well-suited to the diverse experiences of this population. There will be intentionally allowed variation in the dosage of the intervention, which can comprise 6 to 12 sessions, inclusive of an orientation and closing session. The exact number of sessions will be implemented according to the presentation of symptoms and difficulties of the participant. For instance, if the participant presents depressive symptoms concurrent with PTSD, then, the module for the treatment of depression will be included alongside with the module for the treatment of PTSD and the number of sessions will be higher. COMPASS also offers modules in emotion regulation, problem solving, and migration adaption skills which can be added, if they are identified as relevant for the specific participant. For instance, if the participant presents with concrete problems (e.g. precarious housing conditions, or lack of and need of childcare support), then the “problem-solving” component will be included as part of the intervention and the number of sessions will be higher. Additionally, if a client struggles with gaining mastery of any treatment component, an additional session/s, will be added if needed, to ensure mastery. The first 6 sessions will be intended to treat the most urgent symptoms, and the following sessions will address other aspects relevant to the mental health of the participants.

COMPASS will be provided remotely via weekly 60-minute video calls. As prior work identified that this population has long working days, even on weekends, flexibility in session scheduling will be offered (e.g. Sundays, early in the morning, late at night, etc.). There will be some flexibility with session cancellations and re-scheduling; however, a standard rule of three cancellations in a row, or more than five cancellations in total will result in referral of the participant to other services.

#### 2.4b COMPASS Manual

Both COMPASS guides and participants can refer to the COMPASS manual describing each session, the activities, and the homework. The COMPASS manual was co-designed with forcibly displaced Venezuelan people, experts in mental health in Peru, and ultimately compiled into a manual in collaboration with a Venezuelan migrant run NGO. This material has been written and designed as ‘client facing’, so that both COMPASS guides and participant can look at it at the same time, to ensure ease of use in session. Additionally, a network of organizations active in mental health and migration are included for COMPASS guides to make referrals, both as part of the strategies (e.g. a list of organizations that offer community activities for the strategy “behavioral activation”) as well as in case of increasing severity or suicidality, homicidality or psychosis (e.g. community mental health centers in the participant’s neighborhood, organizations that provide support to the victims of domestic violence). There is also a module provided on safety planning, which includes immediate calls to supervisors, and as needed the trainers and primary investigator.

#### 2.4c Lay person Providers

The lay person providers (n = 10), who are called COMPASS guides in the protocol, have all been trained via an intensive in-person 80-hour training, which was followed by practice groups, the completion of the protocol with practice clients, and continuous weekly supervisions with their peers. COMPASS guides were recruited via desk review mapping of available services in the region and were eligible to become providers if they were over 18 years of age, expressed interest in participation, and had available time to attend the training and follow up supervision and sessions. COMPASS guides were not required to have any prior mental health experience. The training materials were designed, with consultation from experts in layperson trainings, to be user-friendly and client facing, allowing lay counselors to implement the protocol effectively. This adaptability is essential, as it enables COMPASS guides to customize interventions according to the specific cultural and contextual factors affecting forcibly displaced Venezuelan people. The training resources, used with lay counselors, emphasize assessment and the selection of treatment components. Consequently, the components COMPASS utilized can differ based on the client’s symptomatology, which lay counselors learn to identify through client vignettes in training. They practice making decisions using assessment, clinical presentations, and discussions with supervisors and counselors.

Based on the apprenticeship model^42^, several COMPASS guides were identified as “very competent” in the training to assume the role of COMPASS supervisors. COMPASS supervisors are closely supervised by the COMPASS trainers, who are all professionally trained local counselors, who are in turn supervised by the primary investigator. Supervision occurs weekly and is continuous.

Evaluation of fidelity of COMPASS guides and COMPASS supervisors will take place continuously in supervision. Supervision will occur between COMPASS guides and COMPASS supervisors, COMPASS supervisors and COMPASS trainers, and COMPASS trainers and the rest of the research team. This will provide ample opportunity to assess any risk and the need to enact safety planning, make decisions regarding the intervention components, and to provide timely support in the implementation of the intervention strategies. Additionally, as described later, all sessions will be audio recorded and fidelity will be retrospectively assessed via review of 20% of the recorded sessions with a COMPASS fidelity checklist.

### 2.5 Procedures and Measures

Before screening participants will be contacted by phone by a member of the research staff trained in responsible conduct of research. If potential participants express interest, the researcher will review a written screening consent form on the phone which is administered and signed via RedCap. After completing the screening consent, the research staff member will administer the screening measures. Following screening, eligible participants will complete a written study consent via phone and signed via RedCap, and then will be randomly assigned to the Intervention Group or WLC. Those in the Intervention Group will be eligible to immediately start the COMPASS intervention, and WLC participants will be assigned to wait for 12 weeks. Participant outcomes will be measured via RedCap at baseline (T0), and in three-month intervals first with an endline (T1) following the COMPASS Intervention (Intervention group) or a 12-week wait period (WLC), and then again at 3-month (T2) and 6-month (T3) follow ups (Figure 1). Outcome measures will be systematically assessed at each phase of the study in alignment with the predefined analysis plan (Figure 2). Participants will provide contact information for follow up, including the information of a friend or family member in case of a change of phone number. Research staff will reach out to participants at each eligible time point, even if participants missed prior follow up assessments. If participants decide to withdraw, research staff will follow up to assess reasons for leaving the study. If at any point during screening, intervention, or follow up, it is apparent that participants require a higher level of care they will be referred by their COMPASS guide to services they are eligible for, based on resources compiled by the research team and expert consultation.

**Figure 1.**
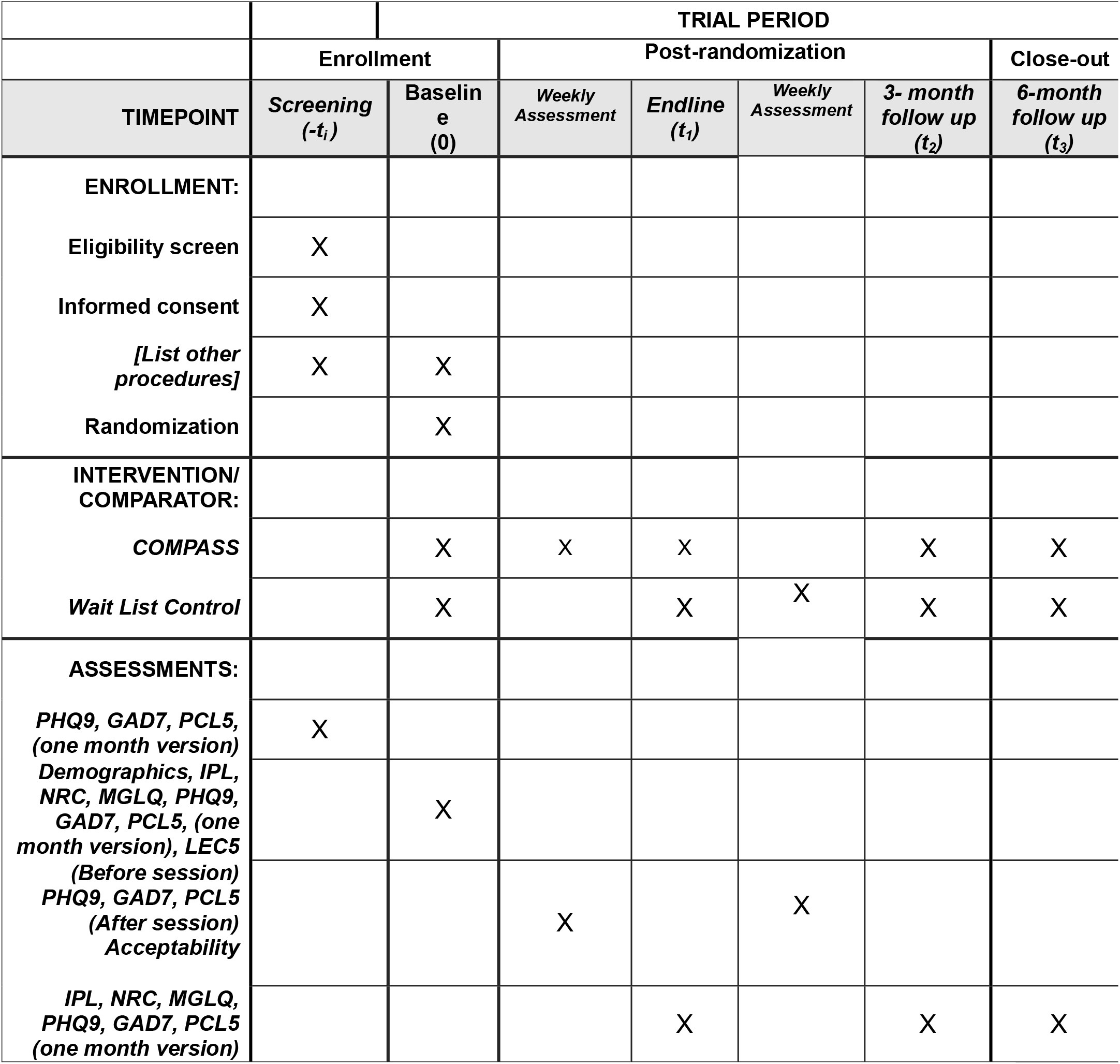
Participant timeline: Schedule of enrollment, interventions, and assessments. Notes: PHQ9 = Patient Health Questionnaire 9, GAD7 = Generalized Anxiety Disorder 7, PCL5 = Post traumatic Stress Disorder Checklist Civilian version 5, IPL = Immigration Policy Lab Integration Index, NRC = Negative Reception Context Scale, MGLQ = Migratory Grief and Loss Questionnaire, LEC5 = Life Events Checklist version 5, COMPASS = Cognitive-behavioral Open-source Mental-health Program Adapted for migrants, Sustainably delivered by lay providers and Supported by evidence Citation: Chan A-W, Boutron I, Hopewell S, Moher D, Schulz KF, et al. SPIRIT 2025 statement: updated guideline for protocols of randomised trials. BMJ 2025;389:e081477. https://dx.doi.org/10.1136/bmj-2024-081477 © 2025 Chan A-W et al. This is an Open Access article distributed under the terms of the Creative Commons Attribution License (https://creativecommons.org/licenses/by/4.0/), which permits unrestricted use, distribution, and reproduction in any medium, provided the original work is properly cited.

**Figure 2.**
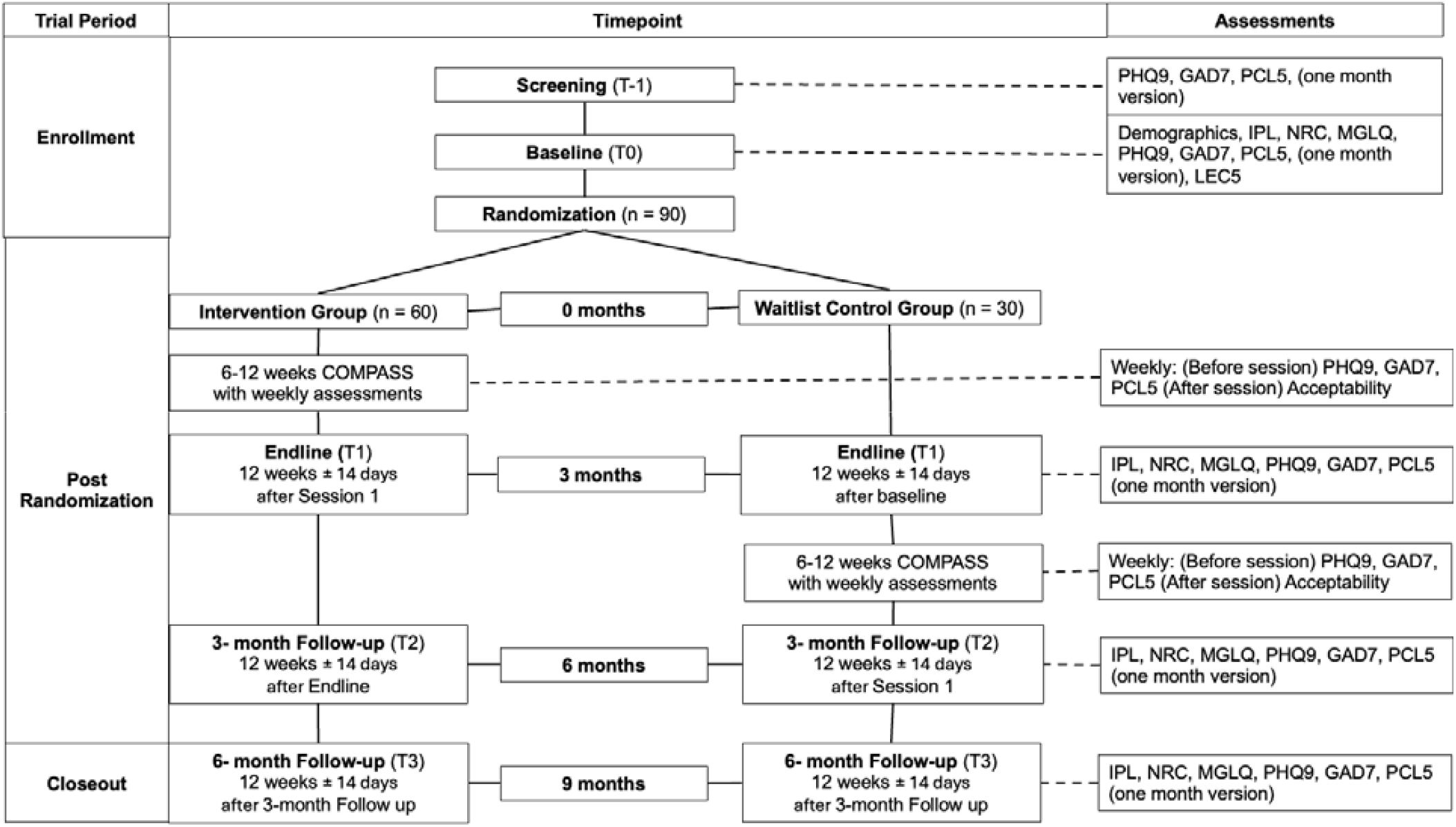
Schematic of Participant timeline: Schedule of enrollment, interventions, and assessments. Notes: PHQ9 = Patient Health Questionnaire 9, GAD7 = Generalized Anxiety Disorder 7, PCL5 = Post traumatic Stress Disorder Checklist Civilian version 5, IPL = Immigration Policy Lab Integration Index, NRC = Negative Reception Context Scale, MGLQ = Migratory Grief and Loss Questionnaire, LEC5 = Life Events Checklist version 5, COMPASS = Cognitive-behavioral Open-source Mental-health Program Adapted for migrants, Sustainably delivered by lay providers and Supported by evidence Citation: Chan A-W, Boutron I, Hopewell S, Moher D, Schulz KF, et al. SPIRIT 2025 statement: updated guideline for protocols of randomised trials. BMJ 2025;389:e081477. https://dx.doi.org/10.1136/bmj-2024-081477 © 2025 Chan A-W et al. This is an Open Access article distributed under the terms of the Creative Commons Attribution License (https://creativecommons.org/licenses/by/4.0/), which permits unrestricted use, distribution, and reproduction in any medium, provided the original work is properly cited.

#### 2.5a Demographic Variables

Demographic variables will be used to assess comparability of the groups. Demographics include age, ethnic and racial background, gender, place of residence, marital status, employment situation, employer, current occupation, highest educational level achieved, and year and month of arrival to Peru.

#### 2.5b Primary outcome measures

The primary outcomes assessed in the study will be symptom changes in measures of common mental health disorders including anxiety, depression, and PTSD. These measures will assess initial effectiveness of the COMPASS intervention. Changes in anxiety will be measured using the Spanish language validated GAD7 measure^34,35^. This is a 7-item instrument that measures generalized anxiety. Changes in depression will be assessed using the Spanish language validated PHQ9 questionnaire^36–38^, which is a 9-item instrument that measures depression. The Spanish language LEC5^40^ which identifies potentially traumatic events in the life course of the participants, will be used at baseline to assess the occurrence of a criterion A event. The Spanish language validated PCL5 will assess symptoms of PTSD^39^. All measures have been validated and used in prior research with this population^8,10,43^.

#### 2.5c Secondary outcome measures

Initial indicators of feasibility and acceptability of the intervention will be assessed as secondary outcomes. Feasibility, or the ability to carry out an intervention in a particular setting, will be assessed via: (a) recruitment rate, (b) retention rate, and (c) fidelity. Recruitment rate will be measured by the percentage of participants who consent to screen for the study, the percentage of participants who meet eligibility for the study, any reasons for ineligibility, the percentage of participants ineligible for each criterion, percentage of eligible participants who enroll, and the characteristics of refusers and reason for refusal. Retention rate will be measured by the number of participants enrolled in the study who complete all study visits and follow up assessments. Fidelity will be measured via recordings of sessions between COMPASS guides and participants. All sessions will be audio recorded, and 20% of the sessions will be randomly selected to be rated 0-10 according to a COMPASS fidelity checklist.

Acceptability, or the perception among guides that the treatment is satisfactory, will be measured via: (a) guide perception, and (b) participant perception. Guide perception will include the guide’s perception of the participant’s satisfaction. After each session, the guides will rate between 0-10 their perceptions of the session’s helpfulness, enjoyment, and relevance for the participant. Additionally, guides will rate their perception (yes/no) on if they believe the participant will use the material learned in that session in the future. Participant perception will be assessed after each session via direct participant report rated from 0-10 on their feelings on that session’s helpfulness, their enjoyment, and the session’s relevance. Additionally, participants will endorse (yes/no) if they plan to use the material learned in the session in the future.

These measures will provide initial information on the feasibility and acceptability of COMPASS for forcibly displaced populations.

#### 2.5d Exploratory outcomes

Exploratory outcomes will include experiences of integration, migratory grief, and perceived negative context of reception. These outcomes will be assessed as moderators to the primary and secondary study outcomes. The Immigration Policy Lab Integration Index-short form (IPL) will assess participant’s experiences with integration^44^. The IPL is a 12-item instrument that measures the psychological, economic, social, navigational, linguistic and political integration in migrant groups. A Peruvian version of the IPL is available and has been used in other research with forcibly displaced Venezuelan people^45^. Migratory grief will be assessed with the Migratory Grief and Loss Questionnaire (MGLQ)^46^. The MGLQ is a 17-item measure validated in Spanish which assesses four factors of migration grief including fear, homesickness, concern and loss of identity. Perceived negative context of reception will be measured with the Perceived Negative Reception Context Scale (NRC)^47^. The NCR utilizes 6 items to assess the degree to which respondents feel unwanted or marginalized within their receiving context. The measure has been used and validated in forcibly displaced Venezuelan populations^47^.

### 2.6 Statistical plan

All analyses will be conducted using multiple packages within R^48^. As this is a pilot study, the primary aim is to compare trends in outcomes between the intervention and WLC groups. Following the intention-to-treat principle, all randomized participants will be included in the analyses. Descriptive statistics will be calculated as frequencies and percentages for categorical variables and as means with standard deviations (or medians with interquartile ranges, as appropriate) for continuous variables. Demographic variables will be examined for differences in distribution using bivariate analyses. Variable distributions will be assessed, and appropriate transformations applied when skewness or violations of parametric assumptions are identified. Reasons for missing data will be systematically tabulated and reported. All quantitative outcomes will be tested at a two-sided significance level of α = 0.05. Regression analyses will be employed to examine changes in primary outcomes, including symptoms of depression, anxiety, and PTSD, as well as initial indicators of feasibility (e.g., recruitment rate, assessment completion rate, attendance, fidelity) and acceptability (e.g., acceptability ratings by therapists and participants). Migration experiences and demographic characteristics will be explored as potential moderators of intervention effectiveness. Regression models will also be used to plot trends in outcome variables across study periods (baseline (T0), endline (T1), and 3-(T2), and 6-month (T3) follow-ups). In addition, descriptive statistics will be calculated to assess participation, retention, fidelity, and satisfaction, and exploratory analyses will investigate associations between covariates, baseline outcome scores, and implementation-related measures to identify characteristics linked to successful delivery of the intervention.

### 2.7 Status and Timeline

As of the writing of this protocol (March 2026), the study is ongoing and in early phases. Recruitment began with the COMPASS guides on April 15 2025. We expect that all study participants will be recruited in November 2026 and data collection complete in April 2027. Analysis is anticipated to be completed by November 2027. The COMPASS guide training, practice groups, and practice participants are complete, with ongoing supervision in progress. Recruitment has started, and the first cohort of participants are finishing their clinical sessions. We expect the last cohort of participants will be completed by February 2027, with results available by April 2028.

### 2.8 Ethical approvals

All procedures for this study were reviewed and approved by ethical review boards at Boston University Medical Center (Reviewed by the Institutional Review Board (IRB) at Boston University Medical Center, Approval number: IRB: H-45280, Approved on: 12/12/24) and Universidad del Pacifico (Reviewed by the Comité de Gestión del Centro de Investigación de la Universidad del Pacífico (CIUP), Approval number: N008-2025/CUIP/UP, Approved on: 1/15/25). Additionally, the study is registered in Clinicaltrials.gov (NCT06635486, submitted 10/7/24, https://clinicaltrials.gov/study/NCT06635486). Any protocol amendments will be submitted for review and approval by the IRB review boards.

### 2.9 Data Management Plan

All survey study data will be collected via the secure online system RedCap, with the research coordinator and Principal Investigator routinely reviewing data quality and addressing issues during weekly investigator meetings. Additional data on the COMPASS guide training, the COMPASS supervisor training, the clinical supervision of COMPASS supervisors by COMPASS trainers, and participant progress will be entered and uploaded in near real time to facilitate monitoring of study operations. To protect confidentiality, participants will be identified only by participant number, visit number, and date of visit, rendering the data de-identified and compliant with the HIPAA Privacy Rule.

## 3. Discussion

### 3.1 Summary

This study seeks to evaluate the initial effectiveness and feasibility of COMPASS, an open-source, lay-delivered CBT program designed to address depression, anxiety, and PTSD. The program was co-developed with forcibly displaced Venezuelan people and mental health experts to ensure cultural and contextual relevance for forcibly displaced Venezuelan people residing in Lima, Peru^11,43^. Previous intervention research with migrant or forcibly displaced populations in Peru and other parts of Latin America has primarily targeted problem-solving skills^49^ or functioned as an entry point within stepped-care models^8^. Thus, COMPASS represents a complementary level of care within task-sharing initiatives, specifically designed to support migration-related adaptation. This study therefore contributes to the growing body of evidence that psychological interventions can be effectively delivered by lay providers, the COMPASS guides, with appropriate modifications, to benefit forcibly displaced populations^8,49^. It also adds to the body of literature which suggests that task-sharing is a promising strategy for resource-constrained settings^5,6,8^. To our knowledge, this is the first study to directly assess the effectiveness, feasibility, and acceptability of a transdiagnostic, open source, lay provided, migration specific intervention of its kind with forcibly displaced Venezuelan people in Lima, Peru.

Preliminary feedback indicates that COMPASS has been well received by guides, participants, and experts. Nonetheless, challenges remain, particularly the absence of a clear referral network and limited stepped-care options to meet the ongoing needs of migrant populations. As part of treatment development, available resources were mapped, revealing a significant gap between high-intensity care and early prevention or low-intensity interventions. To address sustainability, partnerships have been established with multiple NGOs and municipal stakeholders who are committed to supporting the continuation of COMPASS. Findings from this study will provide critical insights into the program’s effectiveness and feasibility in this setting. If COMPASS proves effective, feasible, and acceptable, future research will investigate its sustainability and potential for broader implementation. Ultimately, the approach taken to develop COMPASS, being rooted in evidence based practices^15–20^, implementation science^12,13^, and lived-experience involvement, intentionally builds on existing, effective programs rather than entirely “reinventing the wheel”. The transdiagnostic structure, flexible delivery by trained lay providers, and open-source design of COMPASS position it for adaptation across a wide range of forcibly displaced and migrant populations with minimal modification. Although the current work focuses on forcibly displaced Venezuelan populations in Peru, the model is readily extendable to other Venezuelan communities worldwide, Spanish-speaking migrant groups, and even non-Spanish-speaking forcibly displaced or internally relocated populations within and beyond Peru.

### 3.2 Dissemination plans

A foundational aspect of COMPASS is its open-sourced nature. Guidance in the foundational phase of this research, including from experts in lay person therapies and the Peruvian mental health system, highlight the cruciality of open sourced, low cost, and flexible therapies for scalability and utility for the Peruvian mental health care system, and eventually for generalizability to other health systems. Thus, the study protocol, the statistical methods, the COMPASS manual, training materials, and supervision aides will be made accessible, free of charge on Open Science Framework (OSF) following the completion of this study. The clinical outcomes from the study are sensitive, and there is difficult to completely anonymize. To the extent that we can de-identify data we will make it available in accordance to the National Institute of Mental Health and IRB guidance. Additionally, we plan to publish the results from this study in peer reviewed scholarly journals.

### 3.3 Study Limitations

This study represents the first evaluation of an open-source cognitive behavioral therapy program delivered by lay providers, or the COMPASS guides, to address anxiety, depression, and post-traumatic stress disorder among forcibly displaced Venezuelan people in Lima, Peru. Developed collaboratively with and for this population, the intervention has the potential to substantially expand access to mental health services for forcibly displaced Venezuelan people and, ultimately, other migrant and forcibly displaced communities in Peru and globally. Nonetheless, certain limitations must be acknowledged. In particular, the use of a waitlist control design may overestimate the effectiveness of clinical interventions. However, given the absence of comparable services for this population, this approach represents the most ethical and feasible design for a pilot trial of this nature.

### 3.4 Plans for Amendments or Termination

In the event of adverse events, research personnel will be trained to document detailed descriptions of such occurrences, which will be transmitted electronically to the principal investigator for reporting to the appropriate IRBs within 10 days. At enrollment, participants will receive the contact information for the research coordinator and study site PI to facilitate reporting of adverse events or study-related concerns. A data monitoring committee is not required for the present study, as the intervention is not considered high risk. Monitoring will take place via data collection, clinical supervision, and via direct contact from research participants. Clinical supervision takes place weekly between COMPASS guides and COMPASS supervisors, as well as between COMPASS supervisors and COMPASS trainers. Additionally, COMPASS guides are trained to notify supervisors, who will then contact trainers in the case of any adverse event. Data will be examined annually by the research team for reporting to the sponsor (e.g., preliminary findings, reporting on ethnicity of enrolled participants). Expedited review will be conducted for all events meeting the FDA definition of Serious Adverse Events (SAEs). All SAEs will be reported to BMC and UP IRBs regardless of relatedness to the study, with comprehensive information provided on the event, outcome, concomitant medications, participant medical history and conditions, and relevant laboratory data. Notification of all related study forms will be sent to the IRBs within 2 days of any SAE, and NIH reporting will follow applicable regulations. If at any point investigators or IRBs determine that risks outweigh potential benefits, they retain the authority to recommend study termination.

## Data Availability

No datasets were generated or analysed during the current study. All relevant data from this study will be made available upon study completion. A foundational aspect of COMPASS is its open-sourced nature. Guidance in the foundational phase of this research, including from experts in lay person therapies and the Peruvian mental health system, highlight the cruciality of open sourced, low cost, and flexible therapies for scalability and utility for the Peruvian mental health care system, and eventually for generalizability to other health systems. Thus, the COMPASS manual, training materials, and supervision aides will be made accessible, free of charge following the completion of this study. The clinical outcomes from the study are sensitive, and there is difficult to completely anonymize. To the extent that we can de-identify data we will make it available in accordance to the National Institute of Mental Health and IRB guidance.

https://osf.io/t43de/

